# Monday effect on confirmed cases of COVID-19 in Japan^*^

**DOI:** 10.1101/2021.07.20.21260858

**Authors:** Kuninori Nakagawa, Taro Kanatani

## Abstract

We examined the phenomenon of fewer new confirmed cases of COVID-19 on Mondays in Japan, which we refer to as the Monday effect, and reveal the details of this effect. Specifically, we estimated the difference between the number of new positive cases that decreased over the weekend and the number of new confirmed cases that decreased at the beginning of the week. In Japan, prefectures aggregate and announce the number of confirmed daily cases. This analysis allows us to examine whether there is a Monday effect in each prefecture. We show that the Monday effect is due to the decreased number of inspections on the weekend appearing at the beginning of the week due to a time lag. Our results indicate that the administrative system causes delays in some prefectures, and that some prefectures are less likely to conduct screenings on holidays. Our results also suggest that delays generally occur in prefectures with a population of over 2 million. Congestion, Reporting delay, Public health, COVID-19

## 1 Introduction

In this paper, we analyze the problem of reporting lag in the daily data of COVID-19. In Japan, the number of new positive cases reported on Mondays is relatively low. We refer to this phenomenon as the Monday effect. It began to be mentioned widely after the outbreak period, from July to August 2020. For example, in October 2020, *Asahi Shimbun* [1] reported this effect in Tokyo ^1^.

First, we briefly examine the existence of the Monday effect throughout Japan. By observing the national average of daily cases throughout the period, we can determine how the number of new confirmed cases varies by day of the week (see Table 1).

**Table 1:** National average for each day of the week

|  | Mean | Difference |
| --- | --- | --- |
| SUN | 1397.20 | -100.59 |
| MON | 1023.50 | -474.29 |
| TUE | 1396.09 | -101.70 |
| WED | 1626.60 | 128.81 |
| THU | 1700.53 | 202.73 |
| FRI | 1646.39 | 148.60 |
| SAT | 1695.99 | 198.19 |
| Whole | 1497.79 | — |

The last column presents the difference between the average of daily cases for the entire period and the whole mean for the day of the week. The Monday effect was observed on a nationwide scale, as shown in Table 1. Differences in the means of each weekday seem to be recognized as a nationwide phenomenon. Previously, news channels used to report the raw number of daily cases, but they now add a one-week moving average or a comparison with the same day of the previous week.

We determined that the number of confirmed cases was lower on Mondays. As shown in Table 1, the number of positive cases is lower in the first three days of the week and higher on the other days. In this study, we analyze both the decline in new cases over weekends and reporting lags using daily data announced by each prefecture.

We describe the structure of the decrease in the number of new positive cases reported on Mondays through the lag over the weekend. Many hospitals and public agencies are closed on Saturdays and Sundays. Therefore, the number of inspections is likely to be reduced during weekends. In addition, the closure of public agencies leads to delays in the process of aggregating the reported number of new positive cases. Thus, we can verify the phenomenon of fewer new confirmed cases on Mondays, which implies a lag structure that explains how the decrease in the number of new confirmed cases over the weekend leads to a decrease in the number of new confirmed cases on Mondays. If no lag is detected, the daily release reflects the current inspection result without a delay. We focus on weekends and the two subsequent days, thus examining the effects of Saturday, Sunday, Monday, and Tuesday.

We show that the phenomenon we refer to as the Monday effect in this study can be explained by the structure described above. Regarding the effects within the week, Li (2020)[3] examined time series data on newly confirmed cases of COVID-19 and discussed the weekly recurrence. The author analyzed countries around the world using country-level data and identified an autocorrelation with a 7-day lag.

In Japan, public health management is carried out by the local governments of the 47 prefectures. The central government has delegated the counting of confirmed cases to these local governments. Therefore, the effect is expected to differ for each prefecture. We identified how the structure of time lag differs in each prefecture using daily data on the number of new confirmed cases in each prefecture.

The remainder of this paper is organized as follows. In Section 2, we discuss the data and methods used in the subsequent sections. In Section 3, we first demonstrate the Monday effect in the national-scale data and then present the analysis results for each prefecture. In Section 4, we discuss the implications of our results.

## 2 Method

We describe a time lag between the number of new positive cases that decreased over the weekend and the number of new confirmed cases that decreased at the beginning of the week. Let *x* be the decrease in the number of inspections over the weekend. The decrease in the number of new positives reflected the next day or later is denoted by 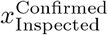. The superscript “Confirmed” indicates the day of the week reflected in the official announcement, and the subscript “Inspected” indicates the day of the inspection. We assume that it takes up to two days for the official reported number of new confirmed cases to reflect *x*_Inspected_. Thus, we have 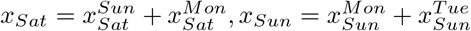.

We provide an example of the impact of a decrease in the number of inspections over the weekend on the decrease in the number of confirmed cases through a lag under the structure currently assumed. We assume that there would be even fewer inspections on Sundays than on Saturdays. We give as an example *x*_*Sat*_ = −10, *x*_*Sun*_ = −20. Without delay, we obtain 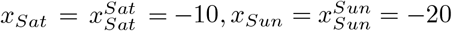. We call this **the no delay**.

Next, we define **the one-day delay** as the delay in reporting ends in one day, with two examples provided. Suppose that 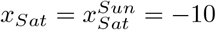 and 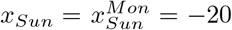, then we have 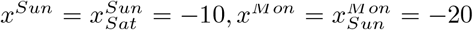. In this example, there will be a decrease in the number of new confirmed cases of −10 on Sunday, −20 on Monday. In another case, suppose that 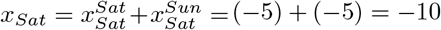 and 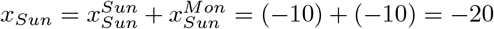, then we have 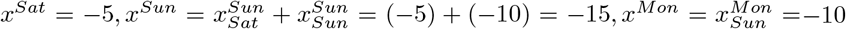. In the latter example, there will be a decrease in the number of new confirmed cases of, 5 on Saturday, 15 on Sunday, and 10 on Monday.

Next, we define **the two-day delay**, where the delay takes two days to end, with an example provided. Suppose that 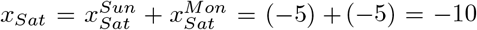 and 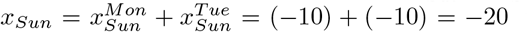 we then have 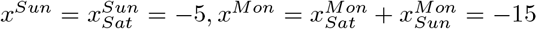, and 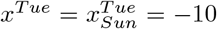. In this example, there will be a decrease in the number of new confirmed cases of 5 on Sunday, 15 on Monday, and 10 on Tuesday. We can also consider an example where reporting delays are concentrated only on Tuesday through the two-day delay.

Moreover, *x*_*Sat*_ = *x*_*Sun*_ = 0 indicates a constant average of inspections within the week. In other words, we cannot identify whether there is a reporting delay. We call this **the constant average**.

We now assume a data-generating process to estimate the delay. It is well known that biological count data tend to be over-dispersed; see Zuur et al. (2010)[9]. To handle such over-dispersion, we assume that the newly confirmed positive cases of the *t*th day, *Y*_*t*_ follow a negative binomial distribution.^2^

In our model, the probability function of *Y*_*t*_ is defined as

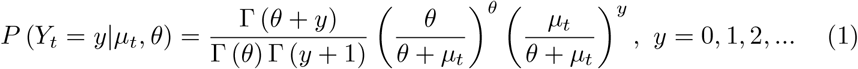

where *µ*_*t*_ is a time-varying deterministic parameter and *θ >* 0.

As the random variable *Y*_*t*_ has the conditional mean and variance

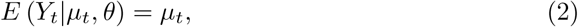

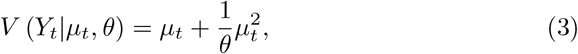

it is clear that *θ >* 0 is the condition of over-dispersion and *θ*^*−*1^ is referred to as an over-dispersion parameter. If *θ* goes to infinity, the distribution converges to the Poisson distribution.

To evaluate the effect of the weekdays, we define the dummy variable

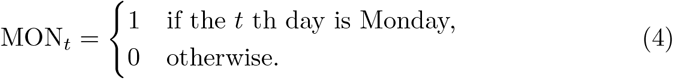

TUE_*t*_, SAT_*t*_, and SUN_*t*_ are similarly defined. We assume that the natural logarithm of *µ*_*t*_ is a linear combination of weekday dummies and a time trend *t*,

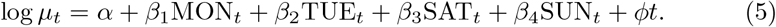

Note that *ϕ* represents the compound daily growth rate of the conditional mean *µ*_*t*_.^3^

As the prefectural data series of daily new cases contain a certain number of zeros in some prefectures, we modify our model to handle such data.

The probability function of zero-inflated negative binomial regression (ZINB) is defined as^4^

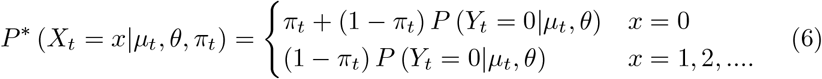

where

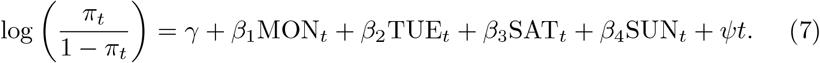

Note that the probability *π*_*t*_ is a time-varying deterministic parameter. We estimate all 12 parameters in (5), (7), and *θ* using the maximum likelihood method,^5^ then select the variables in (5) and (7) via a stepwise approach based on Akaike’s information criterion (AIC). In our approach, we select and delete the same variables in both (5) and (7) to limit the computational complexity.

## 3 Results and Discussion

We use daily data on the number of new positive cases by prefecture obtained from the Nippon Hoso Kyokai (NHK) website[2]. The data spanned 517 days from January 16, 2020, to June 15, 2021, *t* = 1, 2, …, 517.

In subsequent analyses, we identified several patterns in the estimates of the coefficients. We summarize the relationship between the estimates and the delay structures in Table 2. We identify the delays in which the coefficients of weekday dummies are negative and the *p*-value is less than 0.05 (*p <* 0.05).

**Table 2:** Delay Structures

|  | SAT | SUN | MON | TUE |
| --- | --- | --- | --- | --- |
| Without delay 1 | * | * |  |  |
| Without delay 2 |  | * |  |  |
| One-day delay 1 | * | * | * |  |
| One-day delay 2 |  | * | * |  |
| One-day delay 3 |  |  | * |  |
| Two-day delay 1 |  | * | * | * |
| Two-day delay 2 |  |  | * | * |
| Two-day delay 3 |  |  |  | * |
| Constant average | No significantly negative dummy variable. |  |  |  |

We first demonstrate the Monday effect in national-scale data (see Table 3). The column called “count” reports the estimates of coefficients in (5) and *θ*. The column called “zero” also presents estimates of the coefficients in (7).

**Table 3:** Nationwide Result - The one-day delay -

|  | count | zero |
| --- | --- | --- |
| Constant | 4.295***<br>(0.093) | 1.694*<br>(0.684) |
| MON | -0.442***<br>(0.112) | 1.584<br>(1.043) |
| t | 0.009***<br>(0.000) | -0.113***<br>(0.028) |
| Log(theta) | 0.285***<br>(0.059) |  |
| AIC | 7757.841 |  |
| N | 517 |  |
| Significance: *** $\equiv p < 0.001$; ** $\equiv p < 0.01$;<br>* $\equiv p < 0.05$. | | |

**Table 4:** Summary of Delays

|  | N. of Prefs. | Pref. Name |  |  |  |  |  |  |  |  |  |
| --- | --- | --- | --- | --- | --- | --- | --- | --- | --- | --- | --- |
| Without delay 1 | 0 |  |  |  |  |  |  |  |  |  |  |
| Without delay 2 | 1 | Aomori |  |  |  |  |  |  |  |  |  |
| One-day delay 1 | 1 | Niigata |  |  |  |  |  |  |  |  |  |
| One-day delay 2 | 3 | Yamanashi | Shiga | Shimane |  |  |  |  |  |  |  |
| One-day delay 3 | 13 | Miyagi | Tochigi | Saitama | Chiba | Tokyo | Kanagawa | Ishikawa | Gifu | Aichi | Osaka |
|  |  | Hyogo | Nara | Okinawa |  |  |  |  |  |  |  |
| Two-day delay 1 | 1 | Mie |  |  |  |  |  |  |  |  |  |
| Two-day delay 2 | 6 | Fukushima | Ibaraki | Gunma | Shizuoka | Okayama | Fukuoka |  |  |  |  |
| Two-day delay 3 | 1 | Hokkaido |  |  |  |  |  |  |  |  |  |
| Constant Average | 20 | Akita | Yamagata | Toyama | Fukui | Nagano | Kyoto | Wakayama | Tottori | Hiroshima | Yamaguchi |
|  |  | Tokushima | Kagawa | Ehime | Kochi | Saga | Nagasaki | Kumamoto | Oita | Miyazaki | Kagoshima |

As shown in Table 3, we obtain the selected model that contains the following variables: constant term, Monday dummy, and time trend. The results indicate that the number of confirmed cases was significantly lower on Monday. This result shows “the one-day delay 3”: that when the number of inspected cases decreases on Sunday, the number of confirmed cases on Monday also decreases through the lag. On a national scale, no effect on Tuesdays implies that the delay appears to end within a day on average.

We examine the differences among the prefectures. Specifically, we identify how the impact of the weekend affects the decrease in the number of new confirmed cases through the delay. For more detailed results for all prefectures, see Tables 5–12 in the Appendix.

**Table 5:** Monday Effect

|  | Hokkaido |  | Aomori |  | Iwate |  | Miyagi |  | Akita |  | Yamagata |  |
| --- | --- | --- | --- | --- | --- | --- | --- | --- | --- | --- | --- | --- |
|  | count | zero | count | zero | count | zero | count | zero | count | zero | count | zero |
| Constant | 1.334***<br>(0.105) | 297.901<br>(572.020) | -0.318<br>(0.255) | 3.772***<br>(0.439) | -0.934**<br>(0.348) | 7.554***<br>(1.335) | -0.054<br>(0.176) | 2.933***<br>(0.454) | -1.768***<br>(0.434) | 1.353*<br>(0.634) | -0.351<br>(0.246) | 2.724***<br>(0.362) |
| MON | -0.241<br>(0.124) | 51.054<br>(2062.883) |  |  | -0.562*<br>(0.219) | 1.424<br>(0.798) | -0.673***<br>(0.142) | -0.859<br>(0.837) | -0.416<br>(0.344) | 0.820<br>(0.575) |  |  |
| TUE | -0.258*<br>(0.122) | -175.491<br>(332.080) |  |  |  |  |  |  |  |  |  |  |
| SAT | 0.061<br>(0.124) | 21.515<br>(39.746) |  |  | 0.373*<br>(0.187) | 0.872<br>(0.671) |  |  |  |  |  |  |
| SUN | -0.115<br>(0.124) | 32.990<br>(66.174) | -0.569**<br>(0.193) | 0.793<br>(0.455) |  |  | -0.240<br>(0.142) | 0.521<br>(0.569) |  |  |  |  |
| t | 0.009***<br>(0.000) | -10.117<br>(19.534) | 0.007***<br>(0.001) | -0.015***<br>(0.002) | 0.007***<br>(0.001) | -0.029***<br>(0.006) | 0.009***<br>(0.000) | -0.022***<br>(0.003) | 0.008***<br>(0.001) | -0.006***<br>(0.002) | 0.006***<br>(0.001) | -0.011***<br>(0.001) |
| Log(theta) | 0.282***<br>(0.064) |  | 0.357**<br>(0.138) |  | 0.253<br>(0.149) |  | 0.313***<br>(0.083) |  | -0.209<br>(0.282) |  | 0.179<br>(0.147) |  |
| AIC | 4671.711 |  | 1862.261 |  | 1512.188 |  | 3050.775 |  | 1250.497 |  | 1821.216 |  |
| N | 517 |  | 517 |  | 517 |  | 517 |  | 517 |  | 517 |  |

Significance: \*\*\* $\equiv p < 0.001$ ; \*\* $\equiv p < 0.01$ ; \* $\equiv p < 0.05$

**Table 6:** Monday Effect

|  | Fukushima |  | Ibaraki |  | Tochigi |  | Gunma |  | Saitama |  | Chiba |  |
| --- | --- | --- | --- | --- | --- | --- | --- | --- | --- | --- | --- | --- |
|  | count | zero | count | zero | count | zero | count | zero | count | zero | count | zero |
| Constant | -0.457**<br>(0.167) | 2.110***<br>(0.383) | 0.494***<br>(0.138) | 2.678***<br>(0.416) | -0.379<br>(0.201) | 1.610**<br>(0.523) | 0.735***<br>(0.144) | 2.638***<br>(0.381) | 2.015***<br>(0.100) | 2.026***<br>(0.442) | 1.771***<br>(0.107) | 0.784*<br>(0.372) |
| MON | -0.378**<br>(0.129) | 0.612<br>(0.513) | -0.506***<br>(0.124) | -0.412<br>(0.643) | -0.353*<br>(0.162) | 1.011<br>(0.798) | -0.559***<br>(0.132) | 0.873<br>(0.520) | -0.450***<br>(0.108) | 1.230<br>(0.638) | -0.314**<br>(0.117) | 1.433*<br>(0.586) |
| TUE | -0.356**<br>(0.135) | 0.270<br>(0.576) | -0.265*<br>(0.121) | -0.637<br>(0.637) |  |  | -0.331**<br>(0.125) | -0.327<br>(0.562) | -0.202<br>(0.105) | 0.457<br>(0.630) | -0.225*<br>(0.115) | 0.790<br>(0.598) |
| t | 0.008***<br>(0.000) | -0.016***<br>(0.002) | 0.008***<br>(0.000) | -0.023***<br>(0.003) | 0.009***<br>(0.001) | -0.022***<br>(0.004) | 0.007***<br>(0.000) | -0.021***<br>(0.002) | 0.008***<br>(0.000) | -0.037***<br>(0.006) | 0.008***<br>(0.000) | -0.027***<br>(0.004) |
| Log(theta) | 0.703***<br>(0.105) |  | 0.529***<br>(0.084) |  | 0.059<br>(0.085) |  | 0.536***<br>(0.086) |  | 0.568***<br>(0.071) |  | 0.436***<br>(0.071) |  |
| SAT |  |  |  |  | 0.031<br>(0.165) | 2.173**<br>(0.751) |  |  |  |  |  |  |
| SUN |  |  |  |  | -0.277<br>(0.163) | 1.106<br>(0.786) | -0.209<br>(0.129) | 0.850<br>(0.492) |  |  | -0.078<br>(0.116) | 1.207*<br>(0.581) |
| AIC | 2528.228 |  | 3281.452 |  | 2935.642 |  | 3076.639 |  | 4828.492 |  | 4754.340 |  |
| N | 517 |  | 517 |  | 517 |  | 517 |  | 517 |  | 517 |  |

Significance: \*\*\* $\equiv p < 0.001$ ; \*\* $\equiv p < 0.01$ ; \* $\equiv p < 0.05$

**Table 7:** Monday Effect

|  | Tokyo |  |  | Kanagawa |  |  | Niigata |  |  | Toyama |  |  | Ishikawa |  |  | Fukui |  |
| --- | --- | --- | --- | --- | --- | --- | --- | --- | --- | --- | --- | --- | --- | --- | --- | --- | --- |
|  | count | zero |  | count | zero |  | count | zero |  | count | zero |  | count | zero |  | count | zero |
| Constant | 3.635***<br>(0.101) | 2.820***<br>(0.778) |  | 2.013***<br>(0.113) | 3.065**<br>(1.172) |  | -0.802***<br>(0.178) | 1.342***<br>(0.374) |  | 0.544**<br>(0.180) | 1.381***<br>(0.300) |  | 0.720***<br>(0.143) | 1.025**<br>(0.319) |  | 0.448*<br>(0.213) | 1.574***<br>(0.303) |
| MON | -0.529***<br>(0.115) | 3.562**<br>(1.223) |  | -0.651***<br>(0.124) | 1.511<br>(1.757) |  | -0.414**<br>(0.155) | 1.518***<br>(0.450) |  |  |  |  | -0.458**<br>(0.174) | 0.059<br>(0.644) |  |  |  |
| TUE | -0.235*<br>(0.113) | 1.324<br>(1.040) |  | -0.289*<br>(0.120) | 1.230<br>(1.218) |  |  |  |  |  |  |  |  |  |  |  |  |
| SUN | -0.203<br>(0.113) | 1.882<br>(1.068) |  | -0.199<br>(0.122) | 2.695<br>(1.428) |  | -0.339*<br>(0.134) | 0.641<br>(0.478) |  |  |  |  |  |  |  |  |  |
| t | 0.007***<br>(0.000) | -0.111***<br>(0.023) |  | 0.009***<br>(0.000) | -0.118**<br>(0.039) |  | 0.008***<br>(0.000) | -0.011***<br>(0.001) |  | 0.003***<br>(0.000) | -0.010***<br>(0.002) |  | 0.005***<br>(0.000) | -0.012***<br>(0.002) |  | 0.002***<br>(0.001) | -0.008***<br>(0.001) |
| Log(theta) | 0.364***<br>(0.061) |  |  | 0.316***<br>(0.064) |  |  | 0.869***<br>(0.127) |  |  | -0.208<br>(0.145) |  |  | 0.022<br>(0.117) |  |  | -0.063<br>(0.188) |  |
| SAT |  |  |  | 0.041<br>(0.121) | 2.527<br>(1.384) |  | -0.277*<br>(0.128) | -1.304<br>(0.978) |  |  |  |  |  |  |  |  |  |
| AIC | 6315.596 |  |  | 5255.722 |  |  | 2235.143 |  |  | 2162.043 |  |  | 2738.758 |  |  | 1726.660 |  |
| N | 517 |  |  | 517 |  |  | 517 |  |  | 517 |  |  | 517 |  |  | 517 |  |

Significance: \*\*\* $\equiv p < 0.001$ ; \*\* $\equiv p < 0.01$ ; \* $\equiv p < 0.05$

**Table 8:** Monday Effect

|  | Yamanashi |  | Nagano |  | Gifu |  | Shizuoka |  | Aichi |  | Mie |  |
| --- | --- | --- | --- | --- | --- | --- | --- | --- | --- | --- | --- | --- |
|  | count | zero | count | zero | count | zero | count | zero | count | zero | count | zero |
| Constant | -0.559**<br>(0.209) | 0.811<br>(0.428) | -0.320<br>(0.207) | 1.328***<br>(0.379) | 0.431**<br>(0.163) | 1.864***<br>(0.352) | 0.357*<br>(0.169) | 1.862***<br>(0.358) | 1.682***<br>(0.123) | 0.308<br>(0.277) | 0.346*<br>(0.158) | 3.160***<br>(0.441) |
| MON | -0.443*<br>(0.184) | -0.623<br>(0.719) | -0.251<br>(0.175) | 0.647<br>(0.504) | -0.435**<br>(0.157) | 0.092<br>(0.575) | -0.526***<br>(0.145) | 0.951<br>(0.522) | -0.474***<br>(0.130) | 0.164<br>(0.492) | -0.395**<br>(0.135) | 0.425<br>(0.548) |
| TUE | -0.142<br>(0.171) | -1.473<br>(1.127) | -0.313<br>(0.172) | 0.482<br>(0.529) |  |  | -0.316*<br>(0.139) | 0.178<br>(0.563) |  |  | -0.336*<br>(0.134) | 0.750<br>(0.543) |
| SUN | -0.505*<br>(0.198) | -0.243<br>(0.669) |  |  | -0.250<br>(0.159) | 0.580<br>(0.505) | -0.072<br>(0.141) | 1.135*<br>(0.480) |  |  | -0.336*<br>(0.132) | 0.568<br>(0.534) |
| t | 0.007***<br>(0.001) | -0.007***<br>(0.002) | 0.008***<br>(0.001) | -0.012***<br>(0.002) | 0.008***<br>(0.000) | -0.016***<br>(0.002) | 0.008***<br>(0.000) | -0.017***<br>(0.002) | 0.009***<br>(0.000) | -0.013***<br>(0.002) | 0.007***<br>(0.000) | -0.022***<br>(0.002) |
| Log(theta) | 0.247<br>(0.174) |  | 0.090<br>(0.119) |  | 0.082<br>(0.088) |  | 0.364***<br>(0.089) |  | 0.142*<br>(0.072) |  | 0.590***<br>(0.099) |  |
| AIC | 2048.077 |  | 2659.232 |  | 3169.697 |  | 3153.052 |  | 4907.320 |  | 2651.972 |  |
| N | 517 |  | 517 |  | 517 |  | 517 |  | 517 |  | 517 |  |

Significance: \*\*\* $\equiv p < 0.001$ ; \*\* $\equiv p < 0.01$ ; \* $\equiv p < 0.05$

**Table 9:** Monday Effect

|  | Shiga |  | Kyoto |  | Osaka |  | Hyogo |  | Nara |  | Wakayama |  |
| --- | --- | --- | --- | --- | --- | --- | --- | --- | --- | --- | --- | --- |
|  | count | zero | count | zero | count | zero | count | zero | count | zero | count | zero |
| Constant | 0.075<br>(0.144) | 2.394***<br>(0.393) | 1.016***<br>(0.120) | 1.338***<br>(0.335) | 2.509***<br>(0.107) | 1.215***<br>(0.327) | 1.434***<br>(0.109) | 1.305***<br>(0.298) | -0.077<br>(0.151) | 2.068***<br>(0.388) | -0.579**<br>(0.204) | 1.045**<br>(0.403) |
| MON | -0.296*<br>(0.129) | -0.180<br>(0.623) | -0.234<br>(0.126) | 0.486<br>(0.555) | -0.456***<br>(0.122) | 0.652<br>(0.521) | -0.687***<br>(0.123) | 0.235<br>(0.506) | -0.279*<br>(0.135) | 0.013<br>(0.621) |  |  |
| SUN | -0.457***<br>(0.131) | 0.344<br>(0.579) |  |  |  |  |  |  |  |  |  |  |
| t | 0.007***<br>(0.000) | -0.019***<br>(0.002) | 0.008***<br>(0.000) | -0.022***<br>(0.003) | 0.009***<br>(0.000) | -0.025***<br>(0.003) | 0.009***<br>(0.000) | -0.019***<br>(0.002) | 0.009***<br>(0.000) | -0.019***<br>(0.002) | 0.007***<br>(0.001) | -0.011***<br>(0.002) |
| Log(theta) | 0.581***<br>(0.097) |  | 0.317***<br>(0.074) |  | 0.254***<br>(0.067) |  | 0.342***<br>(0.072) |  | 0.385***<br>(0.088) |  | 0.099<br>(0.123) |  |
| AIC | 2742.610 |  | 3898.089 |  | 5547.119 |  | 4520.929 |  | 3028.165 |  | 2279.049 |  |
| N | 517 |  | 517 |  | 517 |  | 517 |  | 517 |  | 517 |  |

Significance: \*\*\* $\equiv p < 0.001$ ; \*\* $\equiv p < 0.01$ ; \* $\equiv p < 0.05$

**Table 10:** Monday Effect

|  | Tottori |  | Shimane |  | Okayama |  | Hiroshima |  | Yamaguchi |  | Tokushima |  |
| --- | --- | --- | --- | --- | --- | --- | --- | --- | --- | --- | --- | --- |
|  | count | zero | count | zero | count | zero | count | zero | count | zero | count | zero |
| Constant | -1.496*<br>(0.611) | 4.000***<br>(0.928) | -1.315*<br>(0.533) | 1.758*<br>(0.712) | -1.676***<br>(0.254) | 1.655**<br>(0.593) | 0.014<br>(0.196) | 2.043***<br>(0.420) | -1.038***<br>(0.258) | 1.579***<br>(0.451) | -1.626***<br>(0.293) | 880.953<br>(1063.892) |
| t | 0.005***<br>(0.001) | -0.017***<br>(0.004) | 0.005***<br>(0.001) | -0.009***<br>(0.002) | 0.012***<br>(0.001) | -0.016***<br>(0.003) | 0.009***<br>(0.001) | -0.020***<br>(0.003) | 0.009***<br>(0.001) | -0.012***<br>(0.002) | 0.009***<br>(0.001) | -5.425<br>(6.539) |
| Log(theta) | -0.750***<br>(0.189) |  | -0.606**<br>(0.198) |  | 0.001<br>(0.093) |  | -0.428***<br>(0.080) |  | -0.125<br>(0.124) |  | -0.778***<br>(0.102) |  |
| MON |  |  | -0.890**<br>(0.341) | -1.388<br>(1.649) | -0.366*<br>(0.178) | 0.554<br>(0.849) |  |  |  |  | -0.298<br>(0.270) | 153.574<br>(7487.878) |
| SUN |  |  | 1.754***<br>(0.398) | 1.067<br>(0.564) |  |  |  |  |  |  | -0.488<br>(0.272) | 144.931<br>(8270.120) |
| TUE |  |  |  |  | -0.382*<br>(0.174) | 0.083<br>(0.956) |  |  |  |  | -0.280<br>(0.247) | -685.044<br>(1154.880) |
| SAT |  |  |  |  |  |  |  |  |  |  | -0.192<br>(0.268) | 141.225<br>(11992.200) |
| AIC | 1015.611 |  | 1119.689 |  | 2651.779 |  | 3214.518 |  | 2202.749 |  | 1617.350 |  |
| N | 517 |  | 517 |  | 517 |  | 517 |  | 517 |  | 517 |  |

Significance: \*\*\* $\equiv p < 0.001$ ; \*\* $\equiv p < 0.01$ ; \* $\equiv p < 0.05$

**Table 11:** Monday Effect

|  | Kagawa |  | Ehime |  | Kochi |  | Fukuoka |  | Saga |  | Nagasaki |  |
| --- | --- | --- | --- | --- | --- | --- | --- | --- | --- | --- | --- | --- |
|  | count | zero | count | zero | count | zero | count | zero | count | zero | count | zero |
| Constant | -1.545***<br>(0.305) | 2.393***<br>(0.546) | -0.708*<br>(0.281) | 1.556***<br>(0.398) | -0.187<br>(0.272) | 1.711***<br>(0.369) | 1.903***<br>(0.136) | 10.528***<br>(2.604) | -0.439<br>(0.226) | 1.903***<br>(0.401) | -0.627*<br>(0.317) | 1.977***<br>(0.465) |
| t | 0.009***<br>(0.001) | -0.014***<br>(0.002) | 0.008***<br>(0.001) | -0.009***<br>(0.001) | 0.005***<br>(0.001) | -0.008***<br>(0.001) | 0.007***<br>(0.000) | -0.164***<br>(0.040) | 0.007***<br>(0.001) | -0.013***<br>(0.002) | 0.008***<br>(0.001) | -0.013***<br>(0.002) |
| Log(theta) | -0.022<br>(0.125) |  | -0.361**<br>(0.140) |  | -0.325*<br>(0.162) |  | -0.204**<br>(0.063) |  | 0.004<br>(0.126) |  | -0.551***<br>(0.143) |  |
| SUN |  |  |  |  | -0.079<br>(0.219) | -1.218<br>(0.725) |  |  | -0.124<br>(0.206) | 1.132*<br>(0.525) |  |  |
| MON |  |  |  |  |  |  | -0.546***<br>(0.153) | -0.155<br>(1.295) | -0.262<br>(0.195) | 0.724<br>(0.537) | -0.320<br>(0.262) | 0.919<br>(0.619) |
| TUE |  |  |  |  |  |  | -0.322*<br>(0.152) | -1.114<br>(1.301) |  |  |  |  |
| AIC | 1847.708 |  | 2081.982 |  | 1865.806 |  | 4523.699 |  | 2123.517 |  | 2156.757 |  |
| N | 517 |  | 517 |  | 517 |  | 517 |  | 517 |  | 517 |  |

Significance: \*\*\* $\equiv p < 0.001$ ; \*\* $\equiv p < 0.01$ ; \* $\equiv p < 0.05$

**Table 12:** Monday Effect

|  | Kumamoto |  | Oita |  | Miyazaki |  | Kagoshima |  | Okinawa |  | Nationwide |  |
| --- | --- | --- | --- | --- | --- | --- | --- | --- | --- | --- | --- | --- |
|  | count | zero | count | zero | count | zero | count | zero | count | zero | count | zero |
| Constant | -0.038<br>(0.226) | 1.865***<br>(0.425) | -0.568*<br>(0.252) | 1.979***<br>(0.373) | 0.804*<br>(0.344) | 3.185***<br>(0.496) | 0.215<br>(0.205) | 4.968***<br>(0.955) | 1.743***<br>(0.117) | 3.331***<br>(0.374) | 4.295***<br>(0.093) | 1.694*<br>(0.684) |
| t | 0.008***<br>(0.001) | -0.017***<br>(0.003) | 0.008***<br>(0.001) | -0.010***<br>(0.001) | 0.004***<br>(0.001) | -0.015***<br>(0.003) | 0.006***<br>(0.001) | -0.032***<br>(0.007) | 0.006***<br>(0.000) | -0.023***<br>(0.002) | 0.009***<br>(0.000) | -0.113***<br>(0.028) |
| Log(theta) | -0.370***<br>(0.086) |  | -0.265*<br>(0.133) |  | -0.689***<br>(0.148) |  | -0.134<br>(0.097) |  | 0.656***<br>(0.078) |  | 0.285***<br>(0.059) |  |
| MON |  |  |  |  |  |  |  |  | -0.521***<br>(0.112) | -0.082<br>(0.459) | -0.442***<br>(0.112) | 1.584<br>(1.043) |
| AIC | 2899.060 |  | 2171.299 |  | 2144.427 |  | 2398.735 |  | 3684.325 |  | 7757.841 |  |
| N | 517 |  | 517 |  | 517 |  | 517 |  | 517 |  | 517 |  |

Significance: \*\*\* $\equiv p < 0.001$ ; \*\* $\equiv p < 0.01$ ; \* $\equiv p < 0.05$

First, we discuss the one-day delay. We found three groups in prefectures where the one-day delay was observed. First one is the one-day delay 3, in which the impact through the lag was observed only on Mondays and not on Tuesdays. These are the 13 prefectures highlighted in light red on the map in Figure 1: Miyagi, Tochigi, Saitama, Chiba, Tokyo, Kanagawa, Ishikawa, Gifu, Aichi, Osaka, Hyogo, Nara, and Okinawa. For these prefectures, we can apply the same interpretation as the trends observed on a nationwide scale. Second is one-day delay 2, where the coefficients on both Sundays and Mondays are significantly negative. These are the three prefectures: Yamanashi, Shiga, and Shimane. Third is one-day delay 1, Niigata.

**Figure 1:**
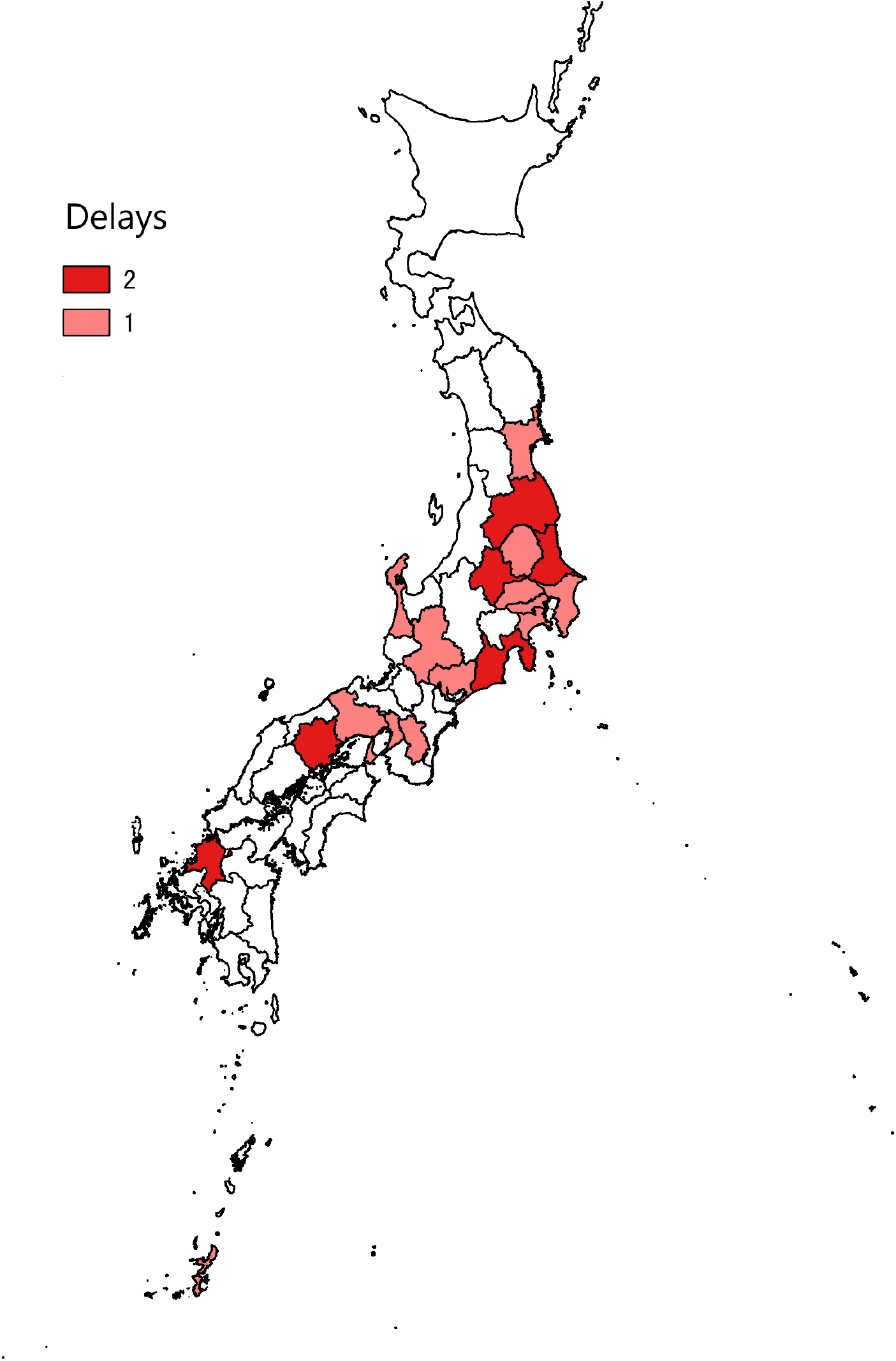
Geographical Distribution of Delay Effects

Second, we discuss six prefectures where the delay was detected on Monday and Tuesday: Fukushima, Ibaraki, Gunma, Shizuoka, Okayama, and Fukuoka. We observed that the reduced number of inspections on the weekend affected both Monday and Tuesday. This delay was the two-day delay 2. These prefectures are highlighted in dark red on the map in Figure 1.

These two results may also indicate that a large population causes a delay in the aggregation of data for official release. The population of each prefecture included in the three groups (the one-day delay 1, 3 and the two-day delay 2) was roughly 2 million or more. This figure is the population threshold set by the Law Concerning the Establishment of Special Wards in Metropolitan Areas (Article 80, 2012). The exceptions are Ishikawa and Okinawa Prefectures, both of which have a population of less than 1.5 million. Furthermore, the three prefectures that consist of the one-day delay 2 group are also small prefectures with a population of less than 2 million.

Third, Hokkaido is the only case of the two-delay 3 where the impact on the weekend is observed only on Tuesday, and all dummy variables remain. However, other dummy variables, except for the TUE dummy variable, are not significant. Hokkaido has a population above 5 million. Additionally, Hokkaido is the largest prefecture in Japan, accounting for 20% of Japan’s total area. The fact that Hokkaido is exceptionally large in both population and size affects the results.

Fourth, we discuss the other prefectures: Aomori, Iwate and Mie. Mie is the only case of the two-delay 1. The result of Aomori indicates that this prefecture reported new positive cases almost without delay, even on weekends. This case is the without delay 2. The number of confirmed cases in Iwate Prefecture was significantly higher on Saturdays and lower on Mondays, a tendency observed only in Iwate. In addition to the operation on weekends, the low number of reported cases of infection itself may have been related to this trend in these prefectures.

Finally, we discuss the prefectures for which we could not obtain any significant dummy variable, that is, the constant average. In our analysis, the constant average corresponds to the case where one of the day-of-week dummies was finally selected. These 20 prefectures are Akita, Yamagata, Toyama, Fukui, Nagano, Kyoto, Wakayama, Tottori, Hiroshima, Yamaguchi, Tokushima, Kagawa, Ehime, Kochi, Saga, Nagasaki, Kumamoto, Oita, Miyazaki, and Kagoshima.

These results are summarised in Table 4 with the exception of the Iwate Prefecture.

Here, we examined whether the conditional mean of confirmed cases was significantly less on Mondays, based on the selected model. We consider it the Monday effect if the coefficient of the MON dummy variable is negative and the *p*-value is less than 0.05 (*p <* 0.05). The Monday effect includes both the one-day delay and the two-day delay. We identified the Monday effect in 25 prefectures. In contrast, we did not find any effect in 22 prefectures.

## 4 Concluding remarks

We estimated the difference between the number of new positive cases that decreased over the weekend and the number of new confirmed cases that decreased at the beginning of the week. We assumed that the number of inspections would be lower on Sundays than on Saturdays. This assumption is consistent with the estimated results. Thus, our results indicate that some prefectures are less likely to conduct screenings on holidays.

We explained that the Monday effect results from delays over the weekend appearing at the beginning of the week with a time lag. The Monday effect was evident in 25 of the 47 prefectures. This effect is related to the areas in which the population is concentrated, as we found that regions with a significant Monday effect include Japan’s three largest metropolitan areas: Tokyo, Osaka, and Nagoya.

Finally, our results suggest a bottleneck in the administrative work of counting the number of new positive cases. In this case, streamlining this process can be performed in advance. This investment can be effective during epidemics/-pandemics.^6^

## Data Availability

All data are obtained from public sources.

https://www3.nhk.or.jp/news/special/coronavirus/

## acknowledgements

We would like to thank Satoru Morita for his helpful comments.

## Footnotes

1 https://www.asahi.com/articles/ASNBY7316NBPUTIL01B.html

2 A quasi-Poisson distribution is also applied for count data with over-dispersion. See Vicuna et al. (2021)[4] for example applications.

3 Chan et al. (2020)[5] estimate a similar growth rate for 18 countries using early-stage data from December 31, 2019, to March 25, 2020.

4 See Benlagha (2020)[6] for an example application of ZINB.

5 We use R[7] software to estimate the ZINB by applying the function `zeroinfl` in the R package `pscl`. See Zeileis et al. (2008)[8] for the application of the R package `pscl`.

6 Regarding the delay in reporting in urban areas, Harris (2020)[10] makes a similar point in the case of New York.

